# BNT162b2 mRNA Vaccine Effectiveness Given Confirmed Exposure; Analysis of Household Members of COVID-19 Patients

**DOI:** 10.1101/2021.06.29.21259579

**Authors:** Sivan Gazit, Barak Mizrahi, Nir Kalkstein, Ami Neuberger, Asaf Peretz, Miri Mizrahi-Reuveni, Tal Patalon

## Abstract

**Importance:** While the mRNA BNT162b2 vaccine effectivness was demonstrated in general population, the question of effectiveness given confirmed exposure has yet been answered, though it has policy implications, as the need for self-quarantine when exposed and protective measures for vaccinated in high-risk areas.

**Objective:** Assessing the BNT162b2 vaccine effectiveness in preventing SARS-CoV-2 infection given high-risk exposure, through analysis of household members of confirmed cases.

**Design:** Retrospective cohort study. Data of household members of confirmed SARS-CoV-2 cases between 20/12/2020 and 17/03/2021 were collected.

**Setting:** Nationally centralized database of Maccabi Healthcare Services (MHS), the second largest Healthcare Maintenance Organization in Israel.

**Participants:** 2.5 million MHS members were considered, of which we included only households with two adult members, given possible lower transmission and susceptibility among children. Households with no prior confirmed infections and a confirmed index case during the study period were included.

**Exposure:** Participants were classified into three vaccination groups in time of the index case (the confirmed exposure)-Unvaccinated; Fully Vaccinated(7 or more days post second dose) and a reference control group of Recently Vaccinated Once(0-7 days from the first dose, presumably still unprotected).

**Main Outcomes and Measures:** Assessing the probability of an additional SARS-CoV-2 infection in the household occurring within 10 days of an index case, calculated separately for the three vaccination groups. Main outcome was vaccine effectiveness given confirmed exposure. High testing rates among household members enabled us to estimate with a high degree of confidence effectiveness against asymptomatic SARS-CoV-2 infection as well.

**Results:** A total of 173,569 households were included, out of which 6,351 households had an index infection (mean [SD] age, 58.9 [13.5] years; 50% were women). Vaccine effectiveness of Fully Vaccinated compared to Unvaccinated participants was 80.0% [95% CI, 73.0-85.1] and 82.0% [95% CI, 75.5-86.7] compared to those Recently Vaccinated Once.

**Conclusion and Relevance:** The BNT162b2 vaccine is effective in a high-risk, real life, exposure scenario, but the protection rates afforded in these settings are lower than those previously described. Household members of COVID-19 patients and any individual with a confirmed exposure to COVID-19 are still at a considerable risk of being infected even if fully vaccinated.

## Introduction

During the COVID-19 pandemic, policymakers are required to make decisions based on incomplete data in an ever-changing environment^1,2^. Any assessment of the effect of the vaccine on community transmission by use of observational data is complicated by two factors. Firstly, the probability of an individual to be infected is greatly influenced by personal behavior and transmission rates in that individual’s immediate surroundings; secondly, the probability of a person to be tested for COVID-19 depends on multiple factors, such as education level or testing availability^3^ – and often on the existence of symptoms.

Using data obtained from the computerized database of the Maccabi Healthcare Services (MHS), the second largest Healthcare Maintenance Organization (HMO) in Israel, we tried to overcome these two potential biases. We analyzed household contacts of confirmed COVID-19 cases, thereby solely including a population at a very high risk of contracting COVID-19^4–7^. Thus, we essentially examined vaccine effectives given confirmed exposure; comparing those exposed (whether in the household or in the community) but not infected to those exposed (whether in the household or in the community) – and infected, across vaccination statuses.

In addition, the probability of a person to be tested for COVID-19 when a household member had already been infected was extremely high in Israel during the study period, therefore allowing for a closer approximative analysis of asymptomatic infection, often not tested and therefore underdiagnosed – potentially impacting the vaccine effectiveness analyses published thus far.

In Israel, a large-scale vaccination program was initiated on December 20, 2020, well before similar efforts were undertaken in other countries^8^. Since within a period of ten weeks more than 50% of the population in Israel was vaccinated with the BNT162b2^9^, data are now available for the analysis of the effectiveness of the vaccine in the prevention of transmission among those at the highest risk of infection, namely household contacts of confirmed COVID-19 cases^10^.

## Methods

We performed an observational cohort study that included household members of confirmed COVID-19 cases, and estimated the effectivness of the mRNA BNT162b2 vaccine vaccine in this high-risk setting. Participants were considered for inclusion from 20/12/2020 onwards, corresponding to the starting date of the national COVID-19 vaccination rollout program. The occurrence of a first episode of infection was recorded up to March 8, 2021; by this time roughly 50% of the entire Israeli population had been vaccinated. Additional household infections were recorded for ten extra days, until March 17, 2021.

### Study population

In Israel all citizens are insured by one of four HMOs as part of a national medical insurance scheme, of which MHS is the second largest with roughly 2.5 million members. For the purpose of this analysis, a household was defined as having two adults. Only households with two adults were included so the protective effect of vaccination could be assessed in a relatively homogenous adult population, given reports of both lower transmission and lower susceptibility in children^12^. Additionally, including more than two members in a household would have required a correction for the number of ‘individuals at risk’ as well as for ‘degree of exposure’. See more in the limitations part of the discussion.

Households with no confirmed COVID-19 infections prior to the study period, with a confirmed index case diagnosed during the study period and with one additional adult who is a member of MHS, were included in the study.

Participants were classified into one of three vaccination status groups in time of the index case (the confirmed exposure): Unvaccinated; Recently Vaccinated Once, i.e. those vaccinated with the first vaccine dose within 0-7 days before the index infection (time of confirmed exposure); and Fully Vaccinated participants, i.e. those who were 7 or more days post the second dose of the vaccine by the time of the confirmed exposure. The second group, i.e. Recently Vaccinated Once, was chosen as a reference period, when the vaccination protective effect is presumably still insignificant (as opposed to persons more than 12 days after the 1^st^ vaccine, when some protection does probably exist)^11^. This enabled us to compare persons who chose to be vaccinated and were either fully protected or not protected at all, and thus to control for possible inherent differences between those who chose not to be vaccinated at all.

COVID-19 was diagnosed with the use of nationally approved SARS-CoV-2 polymerase-chain-reaction (PCR) testing kits.

### Study outcomes

The study’s primary outcome was COVID-19 infection within ten days of COVID-19 diagnosis in an additional adult member of the same household (where an index infection occurred). Both index and additional cases were defined by at least one positive SARS-CoV-2 polymerase chain reaction (PCR) test recorded in the MHS computerized database, considering that all such testing in MHS members is recorded centrally. Household members of confirmed cases were considered not infected if all PCR tests were negative or if no COVID-19 testing was performed.

### Data

The MHS databases were used for this study. As annual disengagement rates are lower than 1%, longitudinal data are available for nearly all persons insured in the MHS. We used existing demographic and background medical data. COVID-19 tests and performance dates were recorded for each patient, as were the dates of the first and second BNT162b2 vaccine administration.

### Statistical analysis

We assessed the probability of a COVID-19 infection occurring up to 10 days after a previously diagnosed COVID-19 infection in a given household.

The probability of being infected was calculated separately for the three vaccination status groups, namely the unvaccinated, those vaccinated within 0-7 days of the first dose of the vaccine, and the fully vaccinated.

For the primary endpoints, vaccine effectiveness was defined for Unvaccinated versus Fully Vaccinated and Unvaccinated versus Recently Vaccinated Once as one minus the risk ratio.

Individuals differ in their decision be tested for COVID-19. Their vaccination status, perceived risk of infection, and symptomatology could all effect this decision^13^. Furthermore, the Israeli Ministry of Health’s regulations indicating that fully vaccinated persons were not obliged to exercise self-quarantine when exposed to a confirmed COVID-19 individual, may have affected the likelihood of vaccinated persons to be tested after an exposure. We hence carried out a secondary analysis attempting to account for differing testing behavior possibly resulting in missed COVID-19 diagnoses. We calculated corrected vaccine effectiveness rates assuming that untested individuals were *as likely* to be infected as tested ones. Therefore, we calculated a corrected effectiveness rate, simulating a scenario in which 100% of adult household members are tested, under the assumption that the probability of test positivity remains constant (rather than decreases in untested individuals). This analysis intends to demonstrate the least-favorable (or strictest) scenario for vaccine effectiveness, enabling us to estimate *lower bound efficiency*.

Our final tier of analyses addressed the problem of defining an “index” infection as opposed to an “additional” infection, in light of a known time lag between the infection event and its detection (positivity lag and symptom lag)^7^. That is, it could be argued that the temporal sequence of household infections reflects the time of *testing* (or *detection*), but not necessarily the time of *infection*, especially when these are close (i.e., the index infection and the additional infection are actually reversed). This could be of importance, as we analyzed the vaccination status of the “additional” infection given confirmed exposure. Therefore, this final analysis examined household members who shared the *same* vaccination status, referred to hereinafter as homogeneously vaccinated couples. This method eliminates the need to determine the order of infections within a given household, as the status of the additional infection is identical to that of the index infection, therefore, the analyzed scenario is one where exposure certainly occurred – and vaccine protectiveness is examined in either the ‘index’ or ‘additional’ household member.

Confidence intervals for binomial probability were calculated, based on Wilson’s score interval. All statistics were performed using Python version 3.1 with the stats models package.

### Ethics declaration

The study protocol was approved by the MHS Institutional Review Board (033-21-MHS). Informed consent was waived by the IRB, as all identifying details of the participants were removed before computational analyses.

## Results

### Participants and rates of additional COVID-19 infections

From December 20, 2020 to March 17, 2021, data were available for 1,312,372 households that included 2,455,924 individuals. We excluded 784,404 households of only one member and 345,365 households with varying numbers of children. Next, we excluded 8,034 households in which an infection was recorded before December 20, 2020.

Out of the remaining 173,569 households, 6,351 households had at least one recorded infection by March 8^th^, 2021. These households were subject to two different methods of analyses (see Methods). The first included 4,024 households stratified into ‘index’ infection and ‘additional’ infection occurring within 10 days across defined vaccination status groups; the second is an analysis of 3,672 homogenously vaccinated couples of the three vaccination groups. (Figure 1).

**Figure 1.**
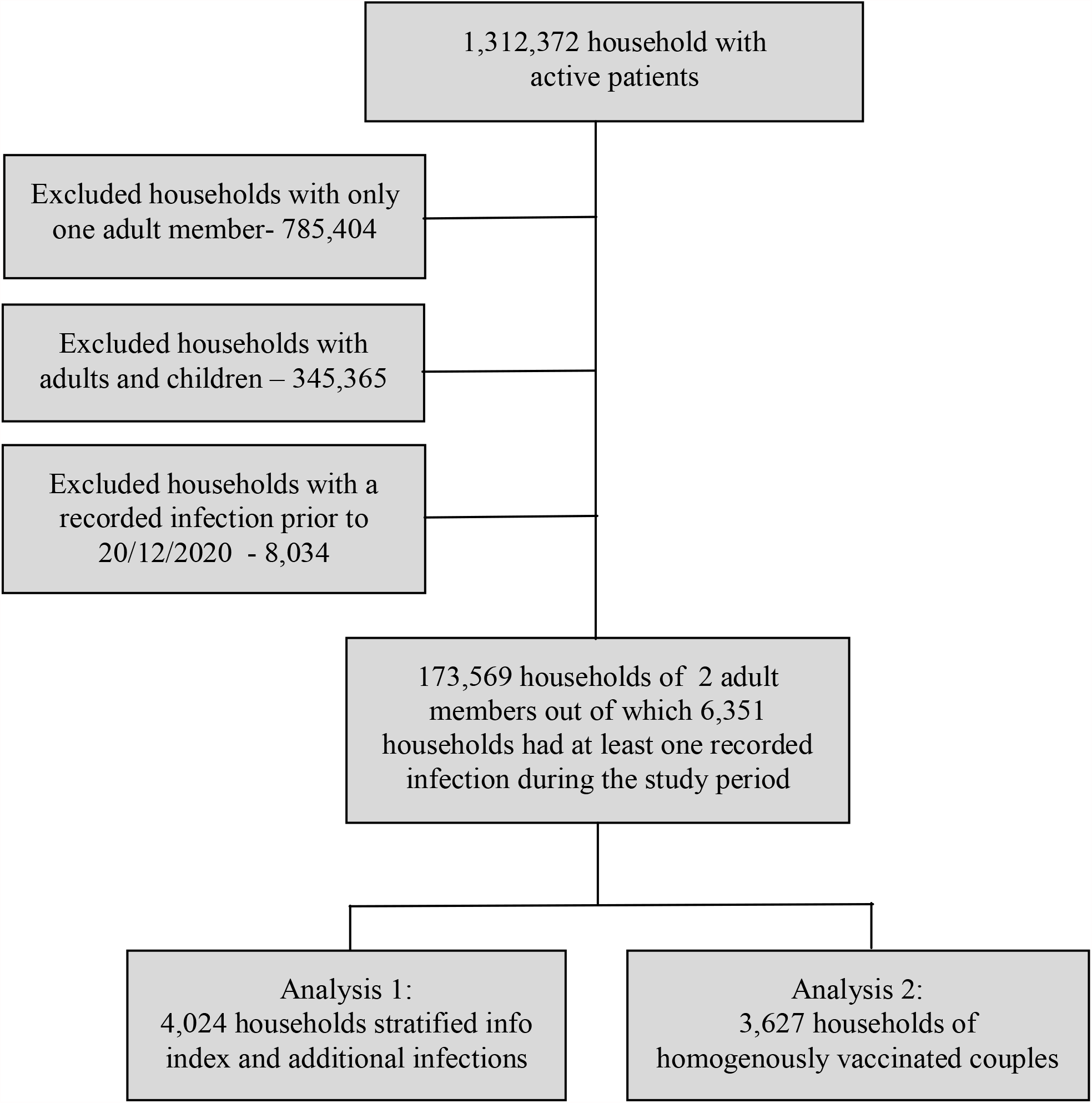
Cohort selection.

Mean age of all adult household members was 58.9 (SD - 13.5), as aged increased with the likelihood of being vaccinated. Females constituted roughly 50% (Table 1). Age and gender of participants by vaccination groups are shown in Supplementary Tables S1 and S3.

**Table 1.** Baseline characteristics of households with a recorded COVID-19 infection

| Characteristic,<br>mean (SD<sup>*</sup>) or % | Households with 2<br>members and at least one<br>recorded infection by<br>March 8<sup>th</sup>, 2020<br>(N = 6,351) | Households stratified into<br>index case and an<br>additional case<br>(N=4,024) | Households of<br>homogenously vaccinated<br>couples<br>(N=3,627) |
| --- | --- | --- | --- |
| Females (%) | 50% | 50% | 50% |
| Mean adult age, years<br>(SD) | 58.9 (13.5) | 57.6 (13.9) | 57.8 (14.5) |
| Households with a second<br>recorded COVID-19<br>infection | 2,729 (43%) | 1,373 (34%) | 1730 (48%) |

\*SD- standard deviation

### Rates of additional household infections occurring 1-10 days after the index infection according to vaccination status groups

We assessed additional infection rates and vaccine effectiveness among household members of infected patients in the three vaccination status groups: Unvaccinated, Recently Vaccinated Once, and Fully Vaccinated.

Rates of additional COVID-19 infections occurring within ten days of the index infection were 37.5% (95% CI-35.7 to 39.3) and 41.7% (95% CI-38 to 45.5) of the Unvaccinated and Recently Vaccinated Once household members of COVID-19 patients, respectively. The proportion of vaccinated household members who tested positive for COVID-19 in the same time period was significantly lower - 7.5% (95% CI-5.6 to 10) (Table 2).

**Table 2.**
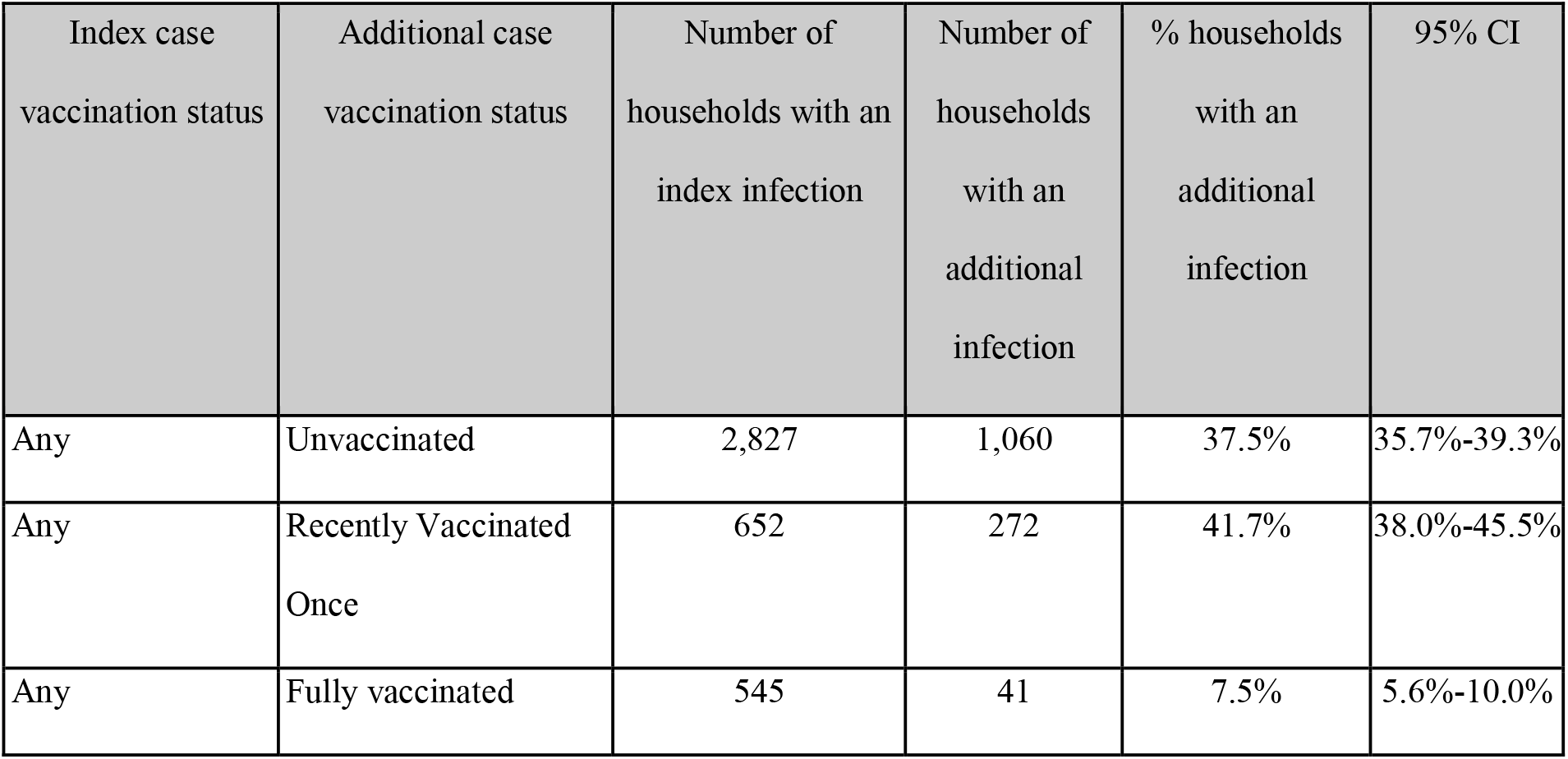
Additional infections occurring 1-10 days after an index infection, by vaccination status

Vaccine effectiveness for Fully Vaccinated compared to Unvaccinated participants was 80.0% (95% CI-73.0 to 85.1), and for Fully Vaccinated individuals compared to those who were Recently Vaccinated Once 82.0% (95% CI-75.5 to 86.7).

### The probability of being tested after being exposed and its effect on estimated vaccine effectiveness

We recorded the likelihood of persons living in households of confirmed COVID-19 cases to be tested for possible infection within ten days of the index diagnosis. The rates of PCR testing were 79.4% (95% CI-77.9-80.9), 85.3% (95% CI-82.3-87.8) and 57.2% (95% CI-53.1-61.3) for the Unvaccinated, Recently Vaccinated Once, and Fully Vaccinated participants. (Figure 2, Supplementary Tables S2).

**Figure 2.**
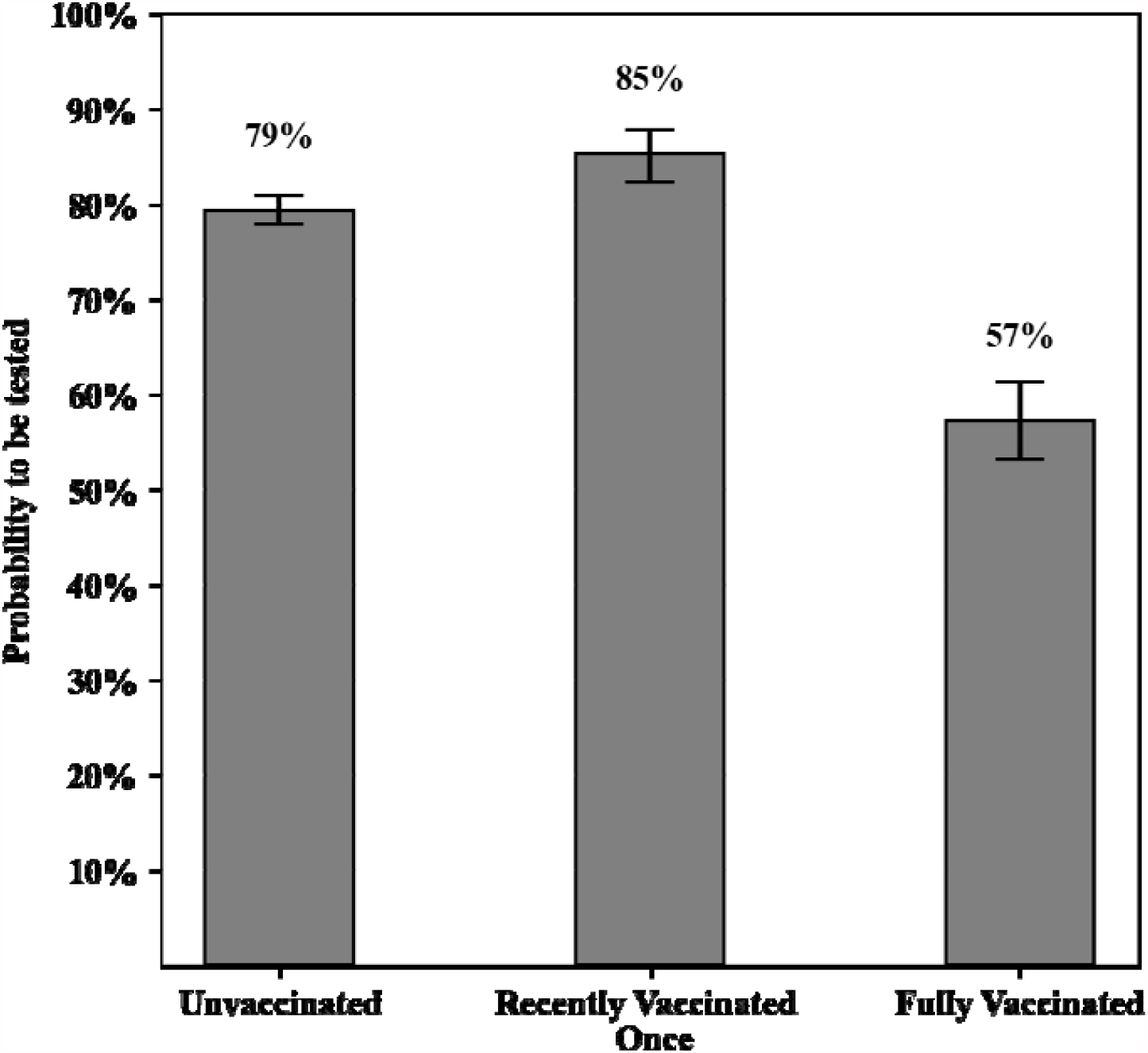
Probability of the second adult in the household to be tested within 10 days of the index infection - cumulative rates of PCR tests among household members of confirmed COVID-19 infections.

As expected, the probability of being tested was lower when the additional household member was a fully vaccinated individual. We calculated corrected effectiveness rates, assuming that untested individuals were *as likely* to be infected as tested ones – thereby estimating *lower bound efficiency* (see methods).

The corrected effectiveness of Fully Vaccinated compared to Unvaccinated individuals was 72.0% (95% CI-65.2 to 77.5) and 73.0% (95% CI-66.0 to 78.5) when Unvaccinated was compared to Recently Vaccinated Once participants.

### Vaccine effectiveness among household members with the same vaccination status (homogeneous couples)

In order to address the problem of defining an “index” infection as opposed to an “additional” infection, we analyzed vaccine effectiveness rates among household members who share the same vaccination status when tested (homogeneously vaccinated couples, see methods).

For each vaccination status groups we calculated the proportion of couples in which both members tested positive within 0-10 days, out of the total number of homogenous couples in which at least one member tested positive.

Additional infections occurred in 49.8% (95% CI-48-51.6) of homogenously unvaccinated couples, in 56.2% (95% CI-51.1-61.1) of homogenous couples recently vaccinated once, and in 12.5% (95% CI-9.1-17.0) of homogenously vaccinated couples within the ten days time window (Table 3). Demographic data of these couples can be seen in Supplementary Table S3.

**Table 3.** Additional infection occurring 0-10 days after an index infection in homogeneously vaccinated couples

| Index case vaccination status | Additional case vaccination status | Number of households with an index infection | Number of households with an additional infection | % households with an additional infection | 95% CI |
| --- | --- | --- | --- | --- | --- |
| Unvaccinated | Unvaccinated | 2975 | 1482 | 49.8% | 48.0%-51.6% |
| Recently vaccinated once | Recently vaccinated once | 381 | 214 | 56.2% | 51.1%-61.1% |
| Fully vaccinated | Fully vaccinated | 271 | 34 | 12.5% | 9.1%-17.0% |

Using this approach, vaccine effectiveness in fully vaccinated individuals was 74.8% (95% CI-65.4 to 81.6) when compared to unvaccinated ones, and 77.7% (95% CI-69.0 to 83.9) when compared to participants recently vaccinated once.

Addressing the probability to be tested by implementing the same method described above yields *lower bound* effectiveness rates, or corrected effectiveness rates. For homogenously vaccinated household members, the corrected effectiveness of fully vaccinated individuals compared to unvaccinated ones was 70.1% (95% CI-61.3 to 76.9%) and 70.5% (95% CI-61.2 to 77.4) compared to the control group of individuals recetly vaccinated once (Supplementary Figure S1 and Supplementary Table S4).

## Discussion

While the mRNA BNT162b2 vaccine effectivness has been well demonstrated in a randomized controlled trial and in retrospective general population studies, the real life vaccine effectiveness in protecting individuals with high-risks of exposure to COVID-19 patients has not been previously demonstrated. Moreover, the question of vaccine effectiveness given confirmed exposure has yet been established, though it derives important policy implications, such as the need for self-quarantine when exposed and lockdown policies in high-risk areas.

In this large cohort, we examined household contacts of confirmed COVID-19 patients, therefore utilizing the microenvironment of the household to best ensure exposure, after which we analyzed vaccine effectiveness. 7.5% of fully vaccinated individuals, who are household members of confirmed COVID-19 patients were infected within ten days. Infection rates among unvaccinated or those who received only one vaccine dose within seven days were much higher at 37.5% and 41.7%, respectively. These figures translate into vaccine effectiveness rates of 80.0% and 82.0% when fully vaccinated persons were compared to participants who were either unvaccinated, or vaccinated only once within seven days of the index infection.

While these rates are somewhat lower than vaccine effectivness rates in the original BNT162b2 randomized controlled trial, and vaccine effectiveness rates observed in a large representative sample of the Israeli population^8^, the degree of protection afforded by the vaccine is still very high, even in this very high-risk exposure scenario. These relatively high effectiveness rates following a high-risk exposure are encouraging news both for the individuals who choose to be vaccinated and for policy makers attempting to decrease viral circulation in the community, especially in light of the difficulty in maintaining social distancing and other protective measures over time.

When we analyzed additional infection rates among households with homogenous vaccination status (i.e. adult household members either both vaccinated or both unvaccinated), we found that in households with two adults who were both fully vaccinated, 12.5% of household members still tested positive for SARS-CoV-2 after exposure. While these rates were much higher for households in which both adults were unvaccinated, the risk of infection for fully vaccinated persons is not negligible and justifies the continuous use of non-vaccine protective measures when fully immunized persons are in contact with COVID-19 patients^15^.

The present study was designed to fill a few gaps in existing data. Firstly, we included persons who were all definitely exposed to SARS-CoV-2 (potentially in the community and definitely in their household), and had an extremely high-risk of being infected. Secondly, we performed secondary analyses correcting for a possible selection bias that stems from the different likelihood of persons to be tested for COVID-19, depending on their vaccination status. In these non-RCT, real-world, scenarios, those not tested are often asymptomatic, or only mildly symptomatic^16–18^. The probability of testing contacts in Israel during the study period were generally very high (79.4%, 85.3% and 57.2% in the three vaccination status groups in this trial). Therefore, although structured screening was not performed, the high testing rates coupled with the correction performed for the differing testing rates in the three study groups, enable us to estimate with a high degree of confidence vaccine effectiveness for COVID-19 that is not necessarily severe and often asymptomatic – thus allowing for a real-world vaccine effectiveness analysis not previously performed.

Our study has several limitations. All data are observational, therefore testing was not performed with the use of a strict clinical protocol. Therefore, testing was less likely to be performed for vaccinated household members and asymptomatic cases may have been missed. We did not include data on viral strains. We assumed that all exposure within a given household posed a high risk of transmission, but we are unable to quantify the actual level of exposure with a given household. The large sample size, high vaccination rates, high testing rates, and correction performed for missing testing all partly overcome these limitations.

Household studies are certainly an efficient way of assessing vaccine effectiveness in a given population^6,19–22^, allowing for analyses of infection rates when exposure is confirmed with a high degree of certainty – thus avoiding the potential skewing effect of intrinsic differences amongst communities. Additionally, there is a high likelihood that the same SARS-CoV-2 strain infected persons living in the same household.

Two issues are the subject of our future research, currently underway; first, the inclusion of more than two household members in general and of children in particular. Transmission dynamics could be influenced by age, and children are more likely to be asymptomatic and less likely to be tested. Children may also affect of the magnitude of exposure as they increase the number of persons per household^12^. “Quantifying” exposure relative to vaccine protectiveness, that is whether being exposed to more than one infected individual can influence vaccine effectiveness, is a complicated question to answer from observational real-world data, which is often incomplete. Secondly, in this study, we have not accounted for the vaccination status of the index case. Our upcoming study attempts to create a model accounting for the possible transmission potential of the index case (rather than solely the susceptibility potential of household members).

## Conclusions

This study shows that the mRNA BNT162b2 vaccine is highly effective in a very high-risk, real life, scenario, but the protection rates afforded in these settings are somewhat lower than those previously described in an RCT or a population-based study. Household members of COVID-19 patients, those caring for them in the healthcare setting, and any other individuals with a confirmed exposure to a COVID-19 patient, are still at risk of being infected^20^ even if fully vaccinated, and should use personal protection measures if possible.

## Data Availability

According to the Israel Ministry of Health regulations, individual-level data cannot be shared openly. Specific requests for remote access to de-identified data should be referred to KSM Research and Innovation Center, Maccabi Healthcare Services.

## Supplementary Material

**Table S1.** Demographic data according to vaccination status in the three vaccination status groups: (N = 4,024)

| Index case vaccination status | Additional case vaccination status | Index case age Mean(SD) | Additional case age Mean(SD) | Index case % female | Additional case % female |
| --- | --- | --- | --- | --- | --- |
| Any | Unvaccinated | 56 (14) | 56 (15) | 49% | 53% |
| Any | Recently vaccinated once | 60 (12) | 61 (11) | 54% | 47% |
| Any | Fully vaccinated | 62 (13) | 63 (10) | 57% | 44% |

\*SD- standard deviation

**Table S2.** Corrected effectivness rates of additional infections within 1-10 days of an index infection, by vaccination status.

| Index case vaccination status | Additional case vaccination status | Number of households | Number of household with testing of the additional (non-index) case | % of household with testing of the additional case | 95% CI | Correction factor to 100% tests of the additional case | Actual number of households with additional infection cases | Actual % of households with additional infection cases | Corrected number of households with additional infection cases | Corrected % of households with additional infection cases | Corrected 95% CI |
| --- | --- | --- | --- | --- | --- | --- | --- | --- | --- | --- | --- |
| Any | Unvaccinated | 2,827 | 2,245 | 79.4% | 77.9%-80.9% | 1.26 | 1,060 | 37.5% | 1,335 | 47.2% | 45.4%-49.1% |
| Any | Recently vaccinated once | 652 | 556 | 85.3% | 82.3%-87.8% | 1.17 | 272 | 41.7% | 319 | 48.9% | 45.1%-52.8% |
| Any | Fully vaccinated | 545 | 312 | 57.2% | 53.1%-61.3% | 1.75 | 41 | 7.5% | 72 | 13.2% | 10.6%-16.3% |

**Table S3.** Demographic data in homogenous couples (N=3,627)

| Index case vaccination status | Additional case vaccination status | Number of households with an index infection | Index case age Mean(SD) | Additional case age Mean(SD) | Index case % female | Additional case % female |
| --- | --- | --- | --- | --- | --- | --- |
| Unvaccinated | Unvaccinated | 2975 | 56 (15) | 56 (15) | 51% | 50% |
| Recently vaccinated once | Recently vaccinated once | 381 | 63 (12) | 63 (12) | 50% | 50% |
| Fully vaccinated | Fully vaccinated | 271 | 68 (9) | 67 (9) | 50% | 50% |

**Table S4.**
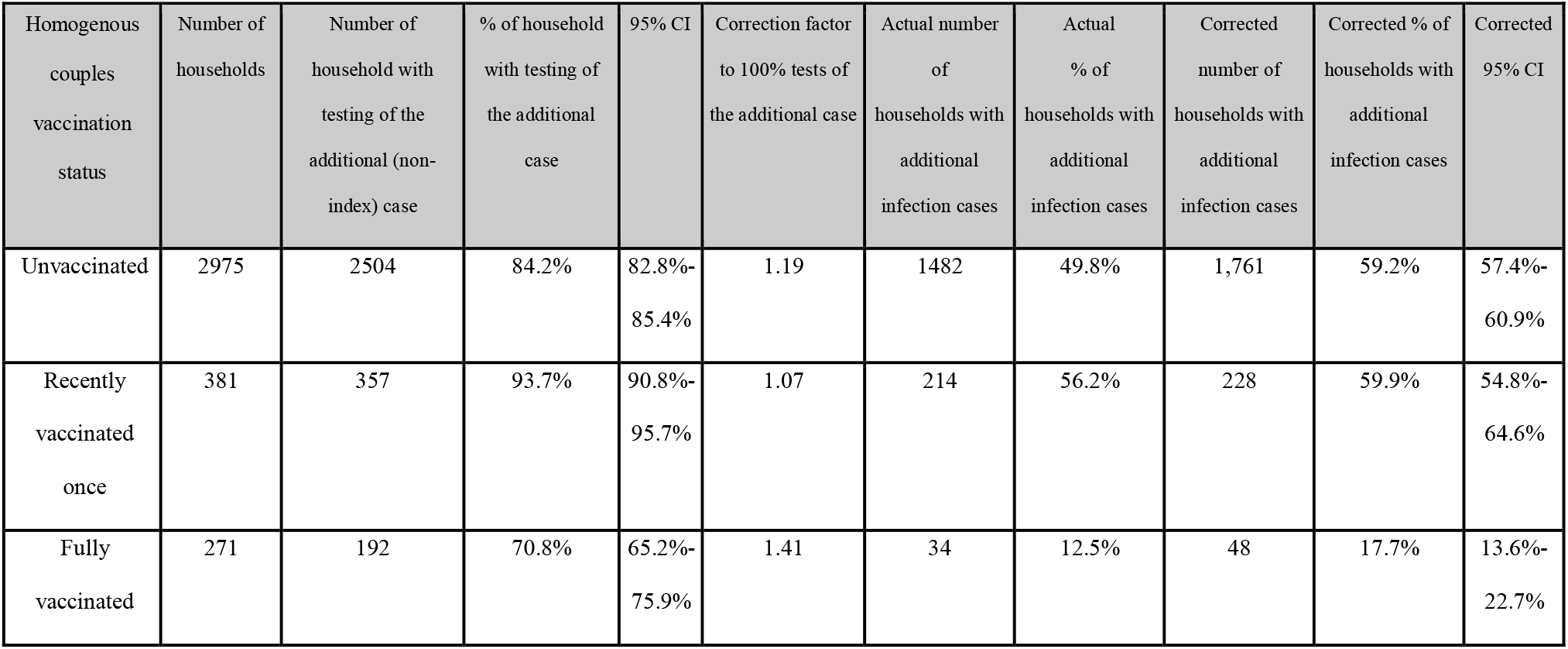
Corrected effectivness rates of additional infections within 0-10 days of an index infection, by vaccination status of homogeneously vaccinated households.

| Homogenous couples vaccination status | Number of households | Number of household with testing of the additional (non-index) case | % of household with testing of the additional case | 95% CI | Correction factor to 100% tests of the additional case | Actual number of households with additional infection cases | Actual % of households with additional infection cases | Corrected number of households with additional infection cases | Corrected % of households with additional infection cases | Corrected 95% CI |
| --- | --- | --- | --- | --- | --- | --- | --- | --- | --- | --- |
| Unvaccinated | 2975 | 2504 | 84.2% | 82.8%-85.4% | 1.19 | 1482 | 49.8% | 1,761 | 59.2% | 57.4%-60.9% |
| Recently vaccinated once | 381 | 357 | 93.7% | 90.8%-95.7% | 1.07 | 214 | 56.2% | 228 | 59.9% | 54.8%-64.6% |
| Fully vaccinated | 271 | 192 | 70.8% | 65.2%-75.9% | 1.41 | 34 | 12.5% | 48 | 17.7% | 13.6%-22.7% |

**Figure S1.**
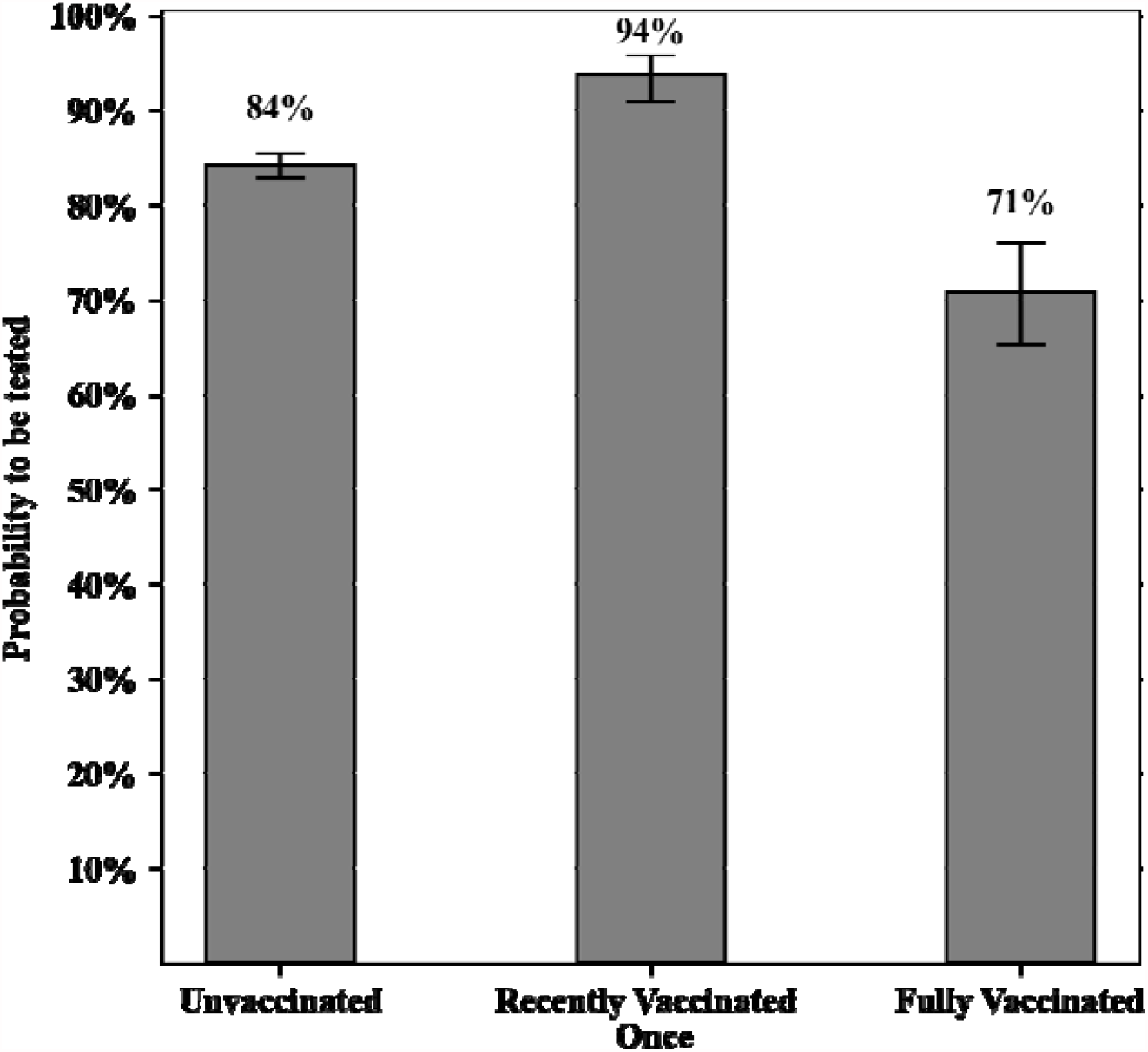
Probability of the second adult in the household to be tested within 10 days of the index infection - cumulative rates of PCR tests among homogenous couples.

